# Development and Validation of Audio-based Guided Imagery and Progressive Muscle Relaxation Tools for Functional Bloating

**DOI:** 10.1101/2022.05.03.22274642

**Authors:** Vincent Tee, Garry Kuan, Yee Cheng Kueh, Nurzulaikha Abdullah, Kamal Sabran, Nashrulhaq Tagiling, Nur-Fazimah Sahran, Tengku Ahmad Iskandar Tengku Alang, Yeong Yeh Lee

## Abstract

Mind-body techniques, including Guided Imagery (GI) or Progressive Muscle Relaxation (PMR), may effectively manage bloating. The current study aimed to develop and validate (psychometric and psychological responses) audio-based GI and PMR techniques for bloating. Audio scripts were first developed from literature reviews and in-depth interviews of participants with bloating diagnosed based on the Rome IV criteria. Scripts were validated using psychometric (content & face validity index) and physiological approaches (brain event-related potentials & heart rate variability). 45/63 participants completed the in-depth interview, and ‘balloon’ emerged as the synonymous imagery description for bloating, of which inflation correlated with a painful sensation. The final tools consisted of narrated audio scripts in the background of a validated choice of music. Overall, content and face validity index for PMR and GI ranged from 0.92 - 1.00. For ERP and HRV, 17/20 participants were analysed. For ERP, there was significant difference between GI and PMR for alpha waves (p=0.029), delta waves (p=0.029) and between PMR and control for delta waves (p=0.014). For HRV, both GI and PMR exhibited similar autonomic responses over controls (overall p<0.05). The newly developed GI and PMR audio-based tools have been validated using psychometric and physiological approaches.

## Introduction

Abdominal bloating is a common problem faced by 76% of patients with disorders of gut-brain interactions (DGBIs) [1–3], also highlighted in a recent global survey [4].

Bloating is commonly associated with a poor quality of life and well-being, affecting productivity, a greater need to seek healthcare, and psychological disturbance [5–10].

Although no treatment is universally regarded as adequate for bloating, several new interventions have been recently developed due to a greater understanding of underlying pathophysiology [11]. The treatment concept is based on the biopsychosocial model and may adopt a multidimensional approach such as the Multidimensional Clinical Profile (MDCP) introduced by the Rome Foundation [12]. Due to central and peripheral-mediated malfunctions in bloating, patients can be treated successfully with behavioural and psychological therapies[13–15]. Experts have recommended mind-body techniques like hypnosis, cognitive behavioural therapy (CBT), and mindful breathing to relieve DGBI-related symptoms [16].

Guided Imagery (GI) is an individualised hypnotic-like mind-body technique commonly employed in sports psychology. The content of GI may include visual, sound, or sensation, and participants are then guided to create a reliable, consistent, and specific content in their minds using their senses. Imagery may be used to transform negative sensations like pain and tension into positive sensations like comfort and relaxation. On the other hand, Progressive Muscle Relaxation (PMR), well-known in the rehabilitative field, is based on tightening and relaxing specific muscle groups to achieve a positive state of relaxation [17,18]. We postulated that GI is superior to PMR in achieving symptom control for bloating. However, to test our hypothesis, the development of a validated intervention is first needed and is a critical component in the current study. Only when there is a validated tool will further randomised trials be feasible. In addition to psychometric validation, the tool can be tested for appropriate physiological changes on the neuronal and autonomic nervous system responses as a form of confirmatory validation.

Hence, the aims of the current study would be first, to develop GI and PMR audio scripts and background music, second, to validate the GI and PMR tools, and third, to determine physiological effects on the brain event-related potentials (ERP) and heart rate variability (HRV).

## Methodology

For the development of audio scripts, extensive literature and established theoretical frameworks were reviewed. The validation of scripts involved content- and face-validity among experts and pilot tested in healthy volunteers. Lastly, for confirmatory validation of the audio scripts, HRV and ERP measurements were determined in a group of healthy volunteers.

### Development of audio scripts

#### Review of literature and theoretical frameworks

For PMR, scripts from published studies were partly adopted to suit local culture and language context[17–20]. For the GI script, relevant studies were adopted to develop content for the in-depth interview[21,22]. In addition, relevant theoretical frameworks were identified to base our GI intervention. These included the Perception Theory in Mind-Body Medicine[23], the Polyvagal theory[24], and the Biopsychosocial model[25]. The Perception Theory in Mind-Body Medicine suggests that mind-body interventions may play a role in realigning perceptual modalities by influencing the cognitive and beliefs system. Meanwhile, the Polyvagal Theory implies bio-mechanisms of phylogenetic vagal responses that affect gut physiological responses. Lastly, the biopsychosocial model reiterates a holistic, multimodal approach based on biology, psychological and social intervention.

#### In-depth interview

After informed consent, participants were invited for in-depth interviews to determine imagery description for bloating. These participants fulfiled the Rome IV diagnostic criteria for functional bloating, were otherwise healthy and did not have any other chronic medical or psychiatric conditions, and on stable medications for their bloating. The sample size was determined based on response saturation. The recorded interviews were transcribed verbatim and analysed thematically by two researchers (VT and NA) using the NVivo software program (QSR International Pty Ltd., Melbourne, Australia). After getting an overview of the general content, further analysis of the text was performed by generating codes, identifying categories, and clustering similar topics[26]. Thereafter, the subcategories would be abstracted into generic and main categories. Codes were cross-checked, and differences were discussed to resolve disagreement, and if not determined, a third reviewer (YCK) would be consulted[27].

#### Narration of GI and PMR scripts

The GI and PMR scripts were written by a health psychologist (GK) and hypnotherapist (VT). The scripts were then discussed, revised and agreed upon by all the investigators in the research team. Subsequently, the scripts were narrated in the local dialect by a local voice-over actor. The voice, tempo, tone, clarity, and enunciation were carefully practised to maximise engagement. Adequate pauses were inserted to allow time and space for listeners. After evaluation, the background music was chosen by seven experts (including one psychiatrist, one gastroenterologist, one health psychologist, and four musicians) on a list of 13 music of different genres. Evaluation was based on the Brunel Music Rating Inventory-2 (BMRI-2)[28], a 6-item questionnaire to assess psychosocial effects of music and the Affect Grid[29] to quantify affect after listening to music. The results then assist a professional music composer (KS) in composing a new background music piece. All audios were recorded and edited using the Logic Pro X software (Apple Inc., USA).

### Validation of audio scripts

#### Content- and face-validity of audio scripts

For content-validity, experts from related fields were invited to rate the audios based on four domains (script content, narrations, experience, and clarity of instructions) using a 4-point Likert scale questionnaire. The rating questionnaire was adapted based on relevant literatures[30–32]. For face-validity, a structured interview session was conducted with healthy volunteers (mean age = 35.57 ± 14.28, 8 men, 24 women). Based on the responses and consensus from investigators, revisions were made to the audio scripts for subsequent pilot testing.

#### Psychometric properties of scripts

The exploratory testing was conducted among healthy volunteers and participants who fulfiled Rome IV-diagnosed functional bloating recruited through purposive sampling after informed consent. Volunteers and patients were excluded in the presence of acute and chronic medical and psychiatric illnesses, hearing impairment, did not have a mobile smartphones and or earphones and recent use of medications which may affect abdominal symptoms. After informed consent, a link containing the audio were sent to their mobile using the WhatsApp application (Facebook Inc., USA). Participants were instructed to listen to the audio when they experienced abdominal bloating. They were told to listen in a quiet, dimly lit room and use an earphone while listening to the audio. At the end of the audio, participants were invited to rate using the same questionnaire for testing validity as above.

#### Physiological responses elicited from listening to audio scripts

Participants with bloating were invited to an air-conditioned dimly lit room with only a maximum of three people were allowed to minimise noise. All electronic devices were turned off to reduce electrical waves interference. With the participant sitting upright on a chair, the MUSE EEG headband version 2016 (InterAxon Inc., Toronto, ON, Canada) (for ERP measurement) and Polar H10 HR sensor chest strap (Polar Electro UK Ltd., Warwick, UK) (for HRV measurement) were placed accordingly. Participants were first asked to listen to a 5-minute audio (control audio) that contained health information on abdominal bloating. Using a cross-over design, participants were then randomised into either the GI or the PMR audio first, then cross-over to the other group in the same session after 15 min interval period.

#### ERP measurements

The MUSE EEG headband has seven sensors with an adjustable band and is worn above the earlobes; three reference sensors (or Fpz) are positioned in the mid-forehead, one anterior frontal-7 (AF-7) sensor in the left forehead, one anterior frontal-8 (AF-8) in the right forehead, two above ear lobe i.e. the left temporal-parietal-9 (TP-9) and the right temporal-parietal-10 (TP-10)[33,34]. These four sensors (AF-7 &8 and TP-9 & 10) would record the brain alpha, beta, gamma, theta, and delta waves (**Supplementary Material 1**). Recorded data would be transmitted to a Mind Monitor mobile app (Mind Monitor, USA) via BlueTooth. Data were cleaned and artefacts (ocular & muscular) were removed using Regression Methods before further analysis [35]. The absolute mean power for each frequency band was calculated by taking the average of four sensors, as shown in **Equation 1** below.

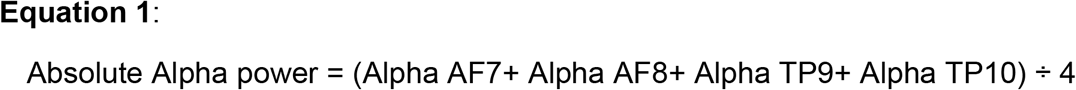

#### HRV measurements

The Polar H10 HR sensor, validated against the ECG gold standard [36], was positioned at the xiphoid process of the sternum using an adjustable chest strap and a Polar M400 monitoring wristband. The recorded R-R interval data was then transferred to the computer using the Polar FlowSync (version 2.5) for analysis. Before processing, the inter-beat intervals (IBIs) were manually corrected for ectopic or missed beats [37]. Kubios HRV software version 3.5 (Biosignal Analysis and Medical Imaging Group, Kuopio, Finland) was used to analyse the parameters. Autoregressive (AR) algorithm was used for power spectral analysis of the frequency series.

#### Data and statistical analysis

Quantitative data were reported as frequency (%) or mean (standard deviation or SD). All analyses were performed with Statistical Package for the Social Science (SPSS) version 26 (IBM Corp., Armonk, NY, USA). Normally distributed values were reported in mean and standard deviation, while median and inter-quartile ranges were reported for non-normally distributed values. Pairwise comparison and repeated measure Analysis of Variance (RM-ANOVA) with Bonferroni adjustment was used for ERP variables, while Wilcoxon-signed rank tests were used for HRV variables to determine the statistical differences in 3 different conditions (GI, PMR and controls), with significant P < 0.05.

## RESULTS

### Development of audio scripts

#### In-depth interview

Of 63 screened participants, 45 patients completed the interview that lasted half an hour (18 did not consent to the study). The themes and examples of quotes from participants for imagery description of bloating are shown in **Table 1**. Three themes emerged, including “balloon” being synonymous with the symptom of bloating, the inflated balloon being the causation of bloating, and the inflated balloon being associated with a sensation of pain or discomfort. All themes were verified based on the quotes. The results of the current interview thus formed the basis of imagery intervention to reduce or deflate the ‘balloon’ in the script as means to improve bloating. Other aspects of the interview included precipitating factors and impact, and the results were addressed in the script as part of the introduction phase to educate participants on bloating.

**Table 1:** Results of Imagery Description of Bloating from the In-depth Interview.

| Domains | Themes | Quotations |
| --- | --- | --- |
| Imagery description of bloating | Bloating as balloon | “I could literally feel my abdomen expand each time I breath...just like a balloon.” (P28, 2020)<br>“My symptoms felt like a balloon in my stomach. It seems to be filled with lots of air and I would burp in order to relieve it.” (P10, 2020)<br>“I think the best description of the bloating symptom would be like having lots of air in the stomach, inflating like a balloon.” (P17, 2020) |
|  | Feeling of inflated balloon | “I felt the bloatedness is like having air in the stomach. I usually felt it at the right side of my abdomen, just below the umbilicus...” (P07, 2020)<br>“The bloating felt like a balloon inflating, always leading to pain and discomfort.” (P16, 2020)<br>“The bloatedness felt like air in the stomach, sometimes it comes with abdominal distention and excessive farts.” (P11, 2020)<br>“The bloating comes unpredictably. It always come together with flatulence, burping and fart. At times, I would feel there is this gas in my stomach, causing my abdomen to become distented.” (P23, 2020)<br>“My bloating felt like having lots of air in my stomach, especially when it’s empty (not eating anything) and I could also feel the abdomen getting bigger.” (P29, 2020) |
|  | Balloon sensation as pain or discomfort | “My bloating sensation comes with a lot of flatulence. I would also feel pain sometimes and it would even lead to me having some difficulties in breathing.” (P19, 2020)<br>“My bloating usually comes with a lot of symptoms. I would feel abdominal pain, breathlessness and even migraine.” (P24, 2020)<br>“I would feel as if there is some form of tightness in my chest, causing me to have difficulty in breathing and sometimes it comes together with abdominal pain.” (P03, 2020)<br>“It was like an expanding balloon and it sounded like a drum when I percussed it (abdomen). Sometimes, it even made me felt breathless, as if the lungs were being pushed upwards by the content.” (P20, 2020) |

### Content of audio scripts

For the GI script, there were six parts, including the following: 1) introduction and instructions, 2) relaxation induction, 3) deepening relaxation and assurance, 4) imagery intervention, 5) gaining control and consolidation, and 6) awakening. For part 1, participants were instructed to accept “unpleasant sensations” that may arise and “be calm and confident” that their unpleasant feelings would “slowly fade away as they become more absorbed in their experience”. Part 2 would facilitate participants into the relaxation state with instructions including “take a deep breath…1,2,3,4,5 and breath it out slowly…1,2,3,4,5. Let all the air in your lungs out”. The third part involved instructions including “for each gentle breath you take…you will drift deeper and deeper into relaxation…” and “you will find better health and a greater sense of control of your symptoms…”. The fourth part would “imagine the balloon in your stomach expands..” and the fifth where participants would gradually gain control over their sensations, making it less intense and bothersome, e.g., “no matter how big the balloons get, you will feel that it does not bother you…and that feeling is going to fade away slowly…”. In the final part, participants are slowly guided to the closure of the session, “in the count of 5, you can slowly focus on your breath…whenever you are ready, you can open your eyes slowly…”.

Similar to the GI script, the first part of PMR was the introduction and instructions, and the relaxation induction phase followed this. The PMR intervention then commenced and involved progressive alternation between contraction and relaxation of different muscle groups, starting from the hands and torso, e.g., “clench your fist as tight as you can…”, to the facial muscles, e.g., “close your eyes as tight as you can and hold it there for 5seconds…”, then neck and shoulder muscles, e.g. “now, focus on your shoulder. Lift your shoulder for 5 seconds…1,2,3,4,5 and relax…”, and finally the abdominal muscle, e.g. “Now, focus on your shoulder. Feel your abdominal muscles and try to breathe in as much as you can. Hold it there for 5 seconds…1,2,3,4,5 and relax…”. The final part was to guide participants to close the session.

### Narration and background music of audio scripts

For the narration, the narrator’s voice was normalised to -10dB and background noises removed. The audio was camouflaged with binaural alpha waves pulse with f =10 Hz. A frequency of 100 Hz was played on the left ear, while 110 Hz was played on the right ear at an acoustic pressure level SPL = 73. For the background music, based on the BMRI-1, the most suitable was music 8 (Indonesian Instrumental) (average score 36), and likewise based on the Affect Grid in the “happy” and “relaxed” grid. Hence, the traditional instrumental music genre was chosen as the theme. The final composed music consisted of traditional Malay instruments (*gambus* and *seruling*) in a major key melody and soundscapes from the natural forest, river flows, and birds. The tempo of the audio was set at 100-120 bpm, equivalent to the average heart rate, to rhythmically entrain and regulate the cardio-respiratory synchronisation while assisting relaxation[38,39].

### Content and face-validity of audio scripts

**Table 2** shows the results of content (CVI) and face validity (FVI) indexes for the four domains (scripts, narrations, audio experience, and clarity). Seven experts, including two staff nurses, two gastroenterology specialists, one health psychologist, and two experienced researchers, were invited for content validity. Overall, experts were generally satisfied, and there were several suggested revisions, including the following: 1) simplifying breathing during relaxation, 2) removing background noises and improving the narrator’s voice, and 3) language. On point 1), the breathing instructions were modified to “breathe in and out calmly and slowly and take the time you need”. On point 2), background noises were filtered using the Logic Pro X software and a deeper voice tone from the narrator to enhance specific keywords like “calm”, “slowly”, and “relax”. For point 3), change to cultural appropriate terms and phrases, for instance, “perlahan” (slowly in standard language) with “*koh-koho*” (local dialect). CVIs for all domains ranged from 0.98 to 1.00 for the GI script and 0.96 to 1.00 for the PMR script **(Table 2)**.

**Table 2:**
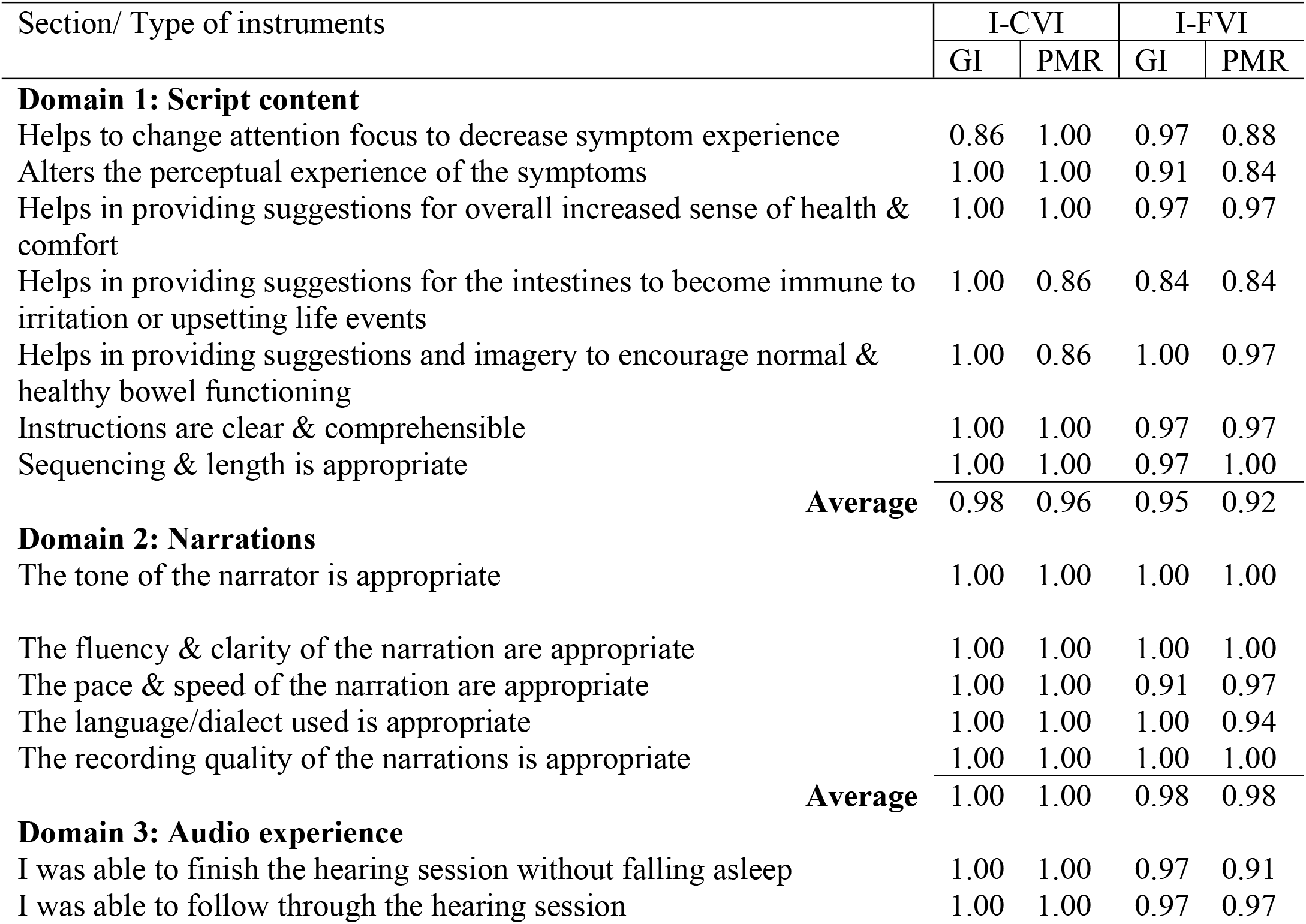
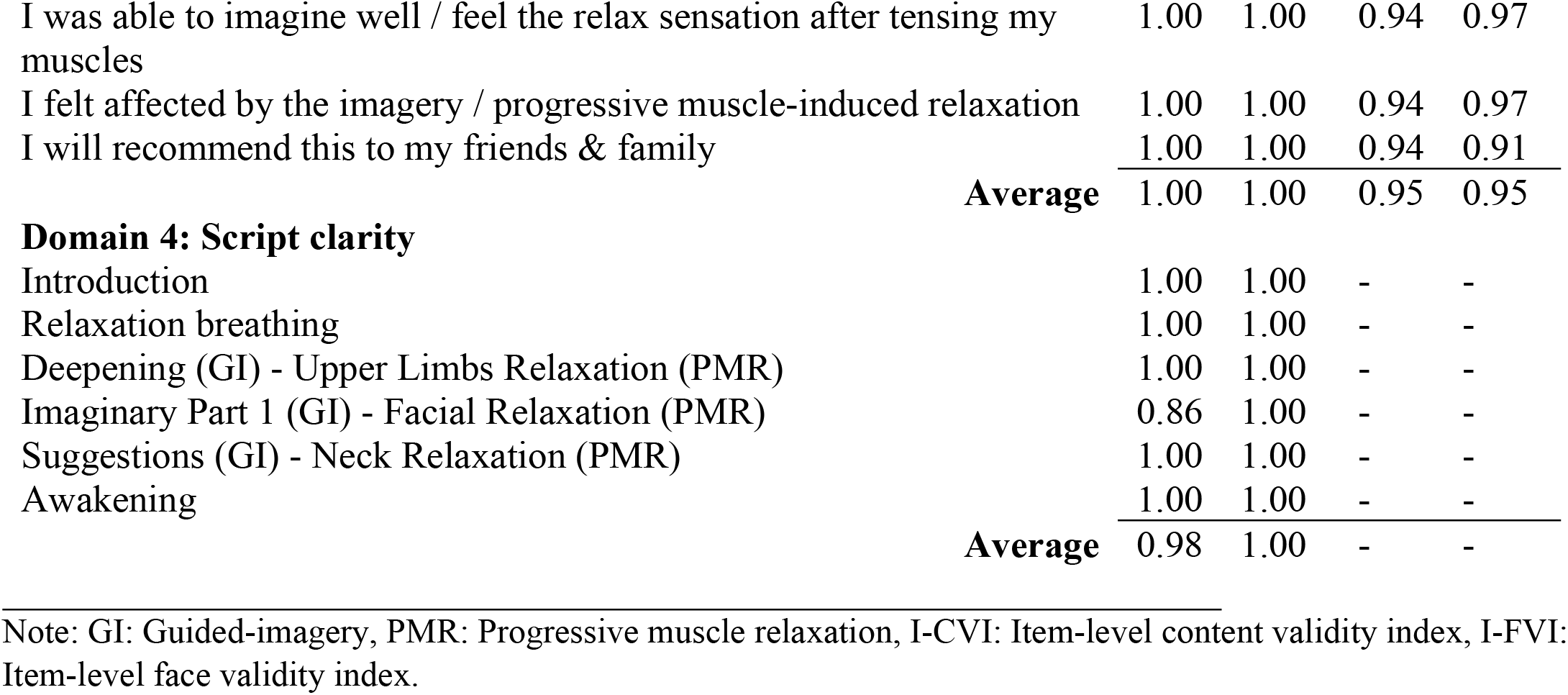
Content and Face Validity of Audio Scripts.

The face-validity from 32 participants with bloating (26 females; mean age 36.7 ± 14.9 years) were likewise positive for both scripts. For example, for the GI script, one participant described it as following: “it’s like magic…I don’t know how but it made me feel really good throughout the day”. Some participants experienced slight discomfort but better after that for example, a participant described the following: “I felt nauseating…after a while, the feeling wear off and I became much comfortable”. There were shortcomings, including 1) unclear narration and background noises at times and 2) difficulties in the balloon imagery. The FVIs for the three domains (all except clarity) ranged from 0.95 to 0.98 for the GI script and 0.92 to 0.98 for the PMR script **(Table 2)**.

### Confirmatory validation based on physiological responses

Of 27 screened healthy volunteers, 20 participants were eventually recruited for the ERP and HRV measurements (7 did not consent to the study). Of the 20, 17 were analysed (eight females, mean age of 23.4±1.7 years), and three were removed from the analysis as they fell asleep. Results are shown in **Figure 2** and **Table 3, respectively**.

**Table 3:**
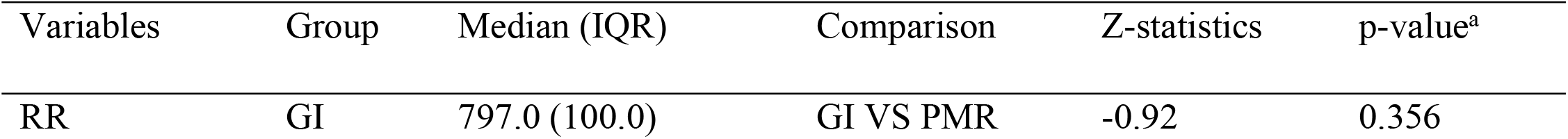
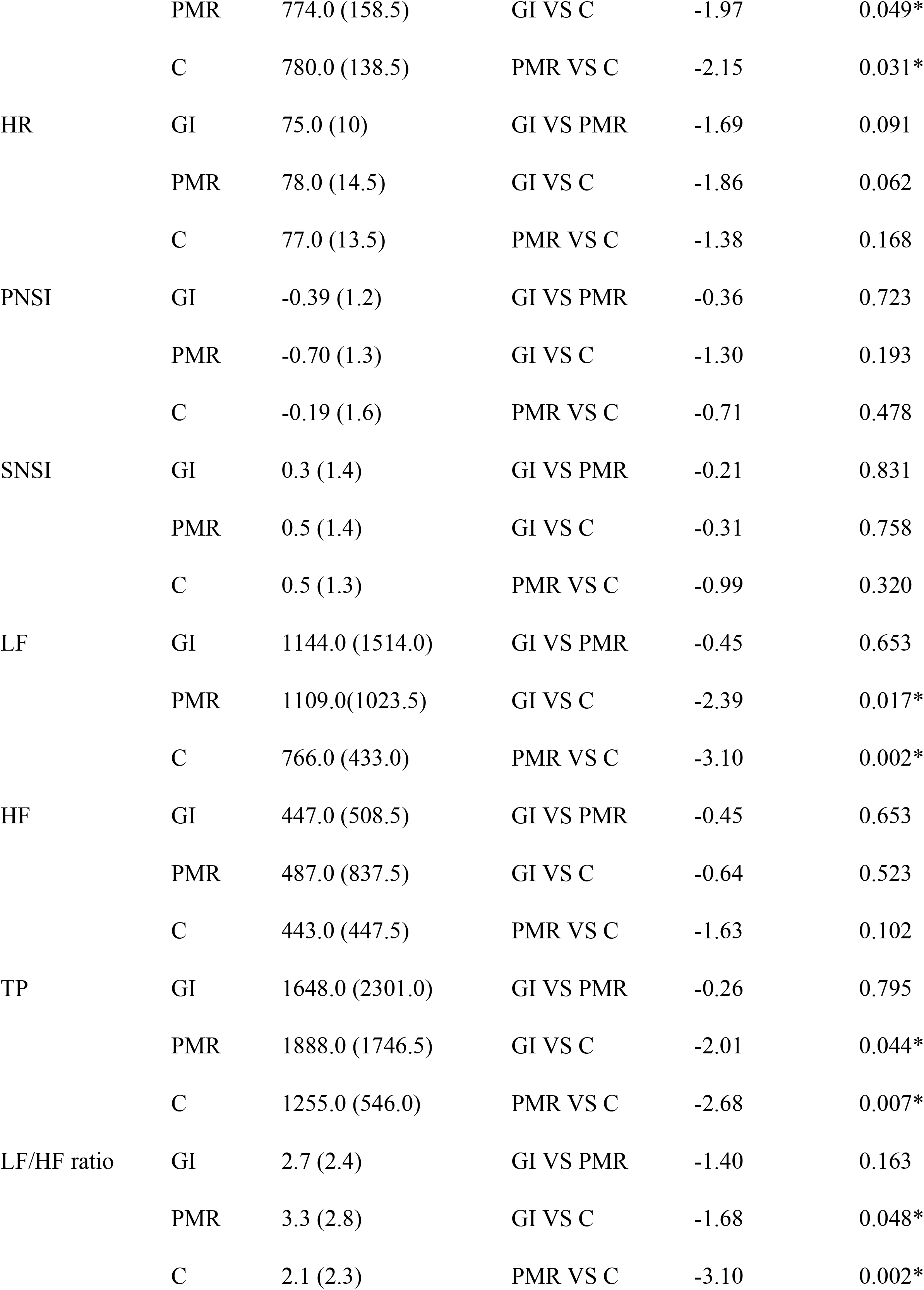
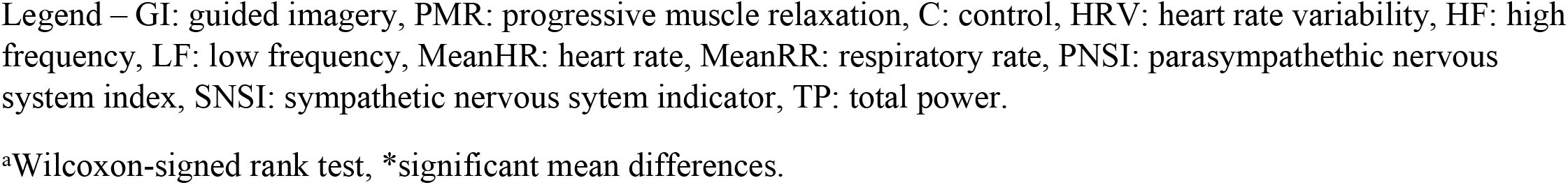
Comparison of Heart Rate Variables between Guided imagery, Progressive Muscle Relaxation and Controls.

Based on the pairwise comparison, ERP analysis revealed a significant mean difference between GI and PMR for alpha waves (p=0.029), delta waves (p=0.029), and between PMR and control for delta waves (p=0.014) only (**Figure 2**). For HRV, using posthoc analysis, significant changes were seen between GI vs. control in the following parameters: RR (p=0.049), LF (p=0.017), TP (p=0.044), and LFHF (p=0.048), and between PMR vs control in the following parameters: RR (p=0.031), LF (p=0.002), TP (p=0.007) and LFHF (p=0.002) **(Table 3)**.

## Discussion

The following is a summary of the main findings: 1) from in-depth interviews and available evidence, bloating, described as ‘balloon’, forms the basis of imagery script, i.e. deflating the ‘balloon’, 2) development of GI and PMR interventions include audio scripts and background music, and demonstrated appropriate content and face validity, 3) there are differences in the ERP and HRV physiological responses between the two interventions, providing confirmatory validation. Overall, the alpha and delta waves differ between GI and PMR, but both GI and PMR have similar autonomic responses.

Development of imagery script involves a thorough literature review, in-depth interview and stepwise modifications by investigators based on comments from various experts. The GI script focuses on ego-strengthening suggestions that emphasise the control of bloating. Bloating as a concept can be interpreted differently across different languages and cultures. The sensation of ‘balloon’, synonymous with bloating, is cross-cultural and allows imagery intervention whereby the balloon can be imagined to ‘deflate’, which will relieve bloating. This aligns with a popular Ericksonian psychotherapy strategy whereby scientific-based therapy was integrated into stories, anecdotes, information, and metaphors [40].

Meanwhile, PMR is a well-known relaxation intervention, and the PMR script focuses on the relaxation of muscle groups around the upper body. Both scripts begin similarly with the phases of introduction and induction (diaphragmatic breathing). Certain words, including “calm” and “slowly” are repetitively mentioned in both scripts to enhance soothing, relaxing effects and to enhance beneficial outcomes from increased exposure [41]. The acceptance of the unpleasant ‘balloon’ sensation in the GI script is essential and is emphasised in the script along with ‘calm’ and ‘slowly’, again to enhance the beneficial outcomes of GI intervention.

The development and validation of scripts were based on a robust methodology as per the recommendation of the Medical Research Council [42]. A robust and validated script would enhance reliability, acceptability, feasibility, and compliance, allowing the retention of research subjects and consistent intervention delivery. In addition to scripts, the background music was incorporated, and music is a common addition to many psychological interventions to enhance therapeutic outcomes further and to facilitate delivery [43]. Again, the choice of music should be culturally appropriate in addition to being relevant to interventions. Our present study employed a validated method to select suitable background music. The process included the use of BMRI-2 scale and the Affect Grid, which allow rating of “emotions” that offer a qualitative and quantitative score, respectively. The complete narrated script with music as a product was then tested for content and face validity. The calculated indexes (CVI and FVI) indicated a robust product.

The products were also tested for effects on brainwaves and autonomic responses as confirmatory validation since a robust intervention should produce appropriate physiological responses. Each brain wave pattern may be affected differently by certain conditions, e.g. meditation enhances theta waves and hypnosis induces alpha waves [44]. It was found in the current study that alpha and delta waves were significantly different between GI vs PMR. Imagery is likened to hypnosis but without affecting consciousness. Thus, it may not be surprising that the alpha wave was different between GI and PMR. Inducing alpha waves could also reduce stress [45] and help depression [46]. Delta wave is usually seen during deep sleep, coma, or anaesthesia [47] and represents comfort and reduced pain [48]. The significant difference with delta waves in GI vs PMR, and PMR vs control, indicated that GI and PMR are interventions that could reduce painful sensations like the ‘balloon’. In addition, recent studies have correlated delta waves with the cognitive process such as attention, problem-solving, and perception [49]. For intervention, especially GI, and to a lesser extent PMR, it involves significant cognitive function to execute a deflation of a balloon or relax the muscles. Background music and bianural beats may also explain the effects seen on brainwaves [50–55]. Various research has shown that music was able to increase alpha and decrease beta waves among individuals with depression [54] or anxiety [39,56].

HRV analysis also confirmed that GI and PMR were different from controls. First, the RRs were significantly different from controls, since GI and PMR required adherence to breathing instructions. Second, low-frequency (LF) power (0.04–0.15 Hz), measuring lower paced respiratory activities on autonomic responses, the LF/HF ratio measuring the sympatho-vagal balance [57,58], and total power measuring overall respiratory responses on autonomic activities were also different from controls. It can be concluded that both GI and PMR induce similar physiological responses to autonomic activities through regulated breathing patterns, which are also an indirect indicator of a relaxation state. For example, Edmond et al. found that an increased LF range correlated with ease and higher comfort level among participants with slow-paced breathing [59]. Similarly, Lin et al. reported an increase in LF power and LF/HF ratio that correlated with an increased perception of relaxation in the paced breathing group [60].

Our study has a few limitations. First, we recognise that different individual has different interpretation and execution of mind-body techniques. This factor may impact the outcome of therapy, especially GI, and we did not screen the imagery ability of participants using a validated tool [61,62]. Second, this study only involved a limited number of participants because of its experimental and exploratory nature. Third, HRV responses might be confounded by other factors besides breathing, including sleep quality which was not covered in our study. Furthermore, HRV responses were conducted while participants were in a ‘relaxed’ state and not while they were ‘stressed’ or experiencing bloating symptoms, which might explain the absence of changes in HR, SNSI and PNSI parameters.

In conclusion, this study’s results indicate that ‘balloon’ that is synonymous with bloating is a suitable imagery tool for intervention, as the imagery of balloon deflation might reduce the symptom. In addition, the study had undertaken a robust methodology in the development and validation of GI and PMR interventions. Besides content and face validity, confirmatory validation of the tools is performed using physiological responses on brainwaves and autonomic responses.

## Data Availability

The minimal data that supports the conclusions of this study are included in the article. The full datasets are not publicly available due to de-identified data sharing restrictions by the Human Research Ethics Committee of Universiti Sains Malaysia. The reason is that the data contain sensitive information (hospital registration number) and are easily identifiable as they come from one single hospital. Data are, however, available from the corresponding author upon reasonable request and with permission of the Human Research Ethics Committee of Universiti Sains Malaysia.

## Acknowledgements

Part of the work herein have been presented at the Asian Pacific Digestive Week (APDW) 2021, 19-22 August 2021 and published as conference-related abstract: Development of cultural specific guided imagery and progressive muscle relaxation therapy for treatment of functional bloating. Journal of Gastroenterology and Hepatology 36 (Suppl. 2), pg156. https://doi:10.1111/jgh.15607. The work has also been presented at the YSN-ASM International Scientific Virtual Conference (ISVC) 2021, 29 March – 1 April and published as conference-related abstract: Development of Guided Imagery and Progressive Muscle Relaxation Therapy Audio for Patients with Functional Bloating. ASM Science Journal 16, pg13. https://doi.org/10.32802/asmscj.2021.isvc. Also, we wish to thank the staff of GI Function & Motility Unit GI of Hospital USM, and other participating investigators for their assistance and input throughout the study.

## Author contributions

YCK, GK, and YYL secured the funding. VT, YCK, GK, NT, KS and YYL contributed to the conception and design of the study. VT, NA, NS, TAITA and YCK collected, analyzed, and interpreted the data. VT is responsible for the manuscript drafting. NT, GK, YCK, and LYY critically revised the paper and enhanced its intellectual content. All authors have reviewed the manuscript before finalizing it into the current version.

## Ethical approval and consent to participate

The study was approved by the Human Research Ethics Committee of Universiti Sains Malaysia (USM/JEPeM/20110562) following the guidelines of the International Declaration of Helsinki. Only consented participants were included in the current study.

## Supporting Information

**S1 Figure.**
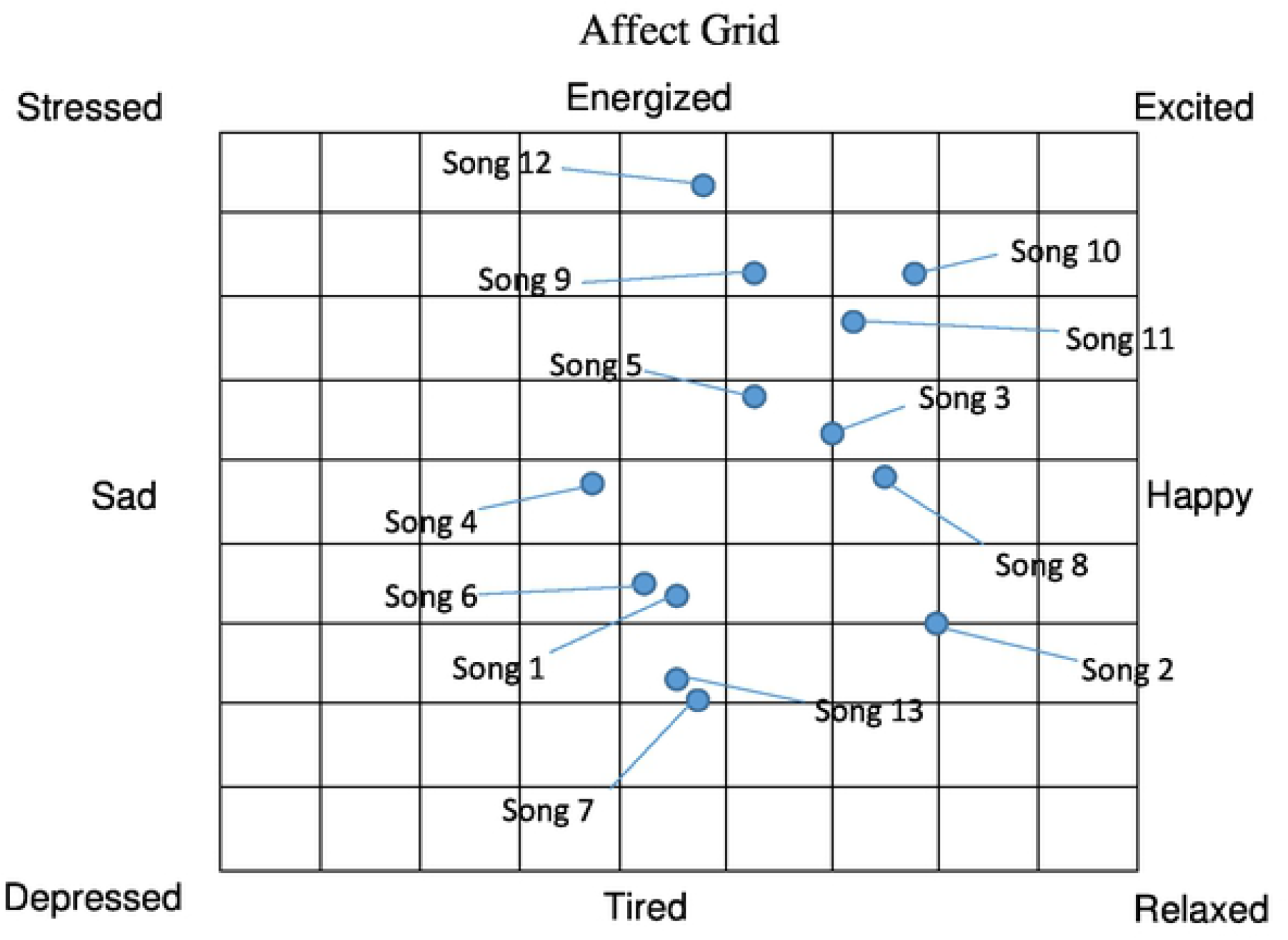
The Affect Grid for the ratings of the 13 selected musical background. Raters were instructed to rate how they are feeling right now and place one checkmark in the grid. The score is calculated according to the number of the squares. The horizontal dimension is used to score the pleasure-displeasure score, counting 1 to 9 starting at the left. The vertical dimension is used to score the arousal-sleepiness score, counting 1 to 9 starting at the bottom.

**S2 Figure.**
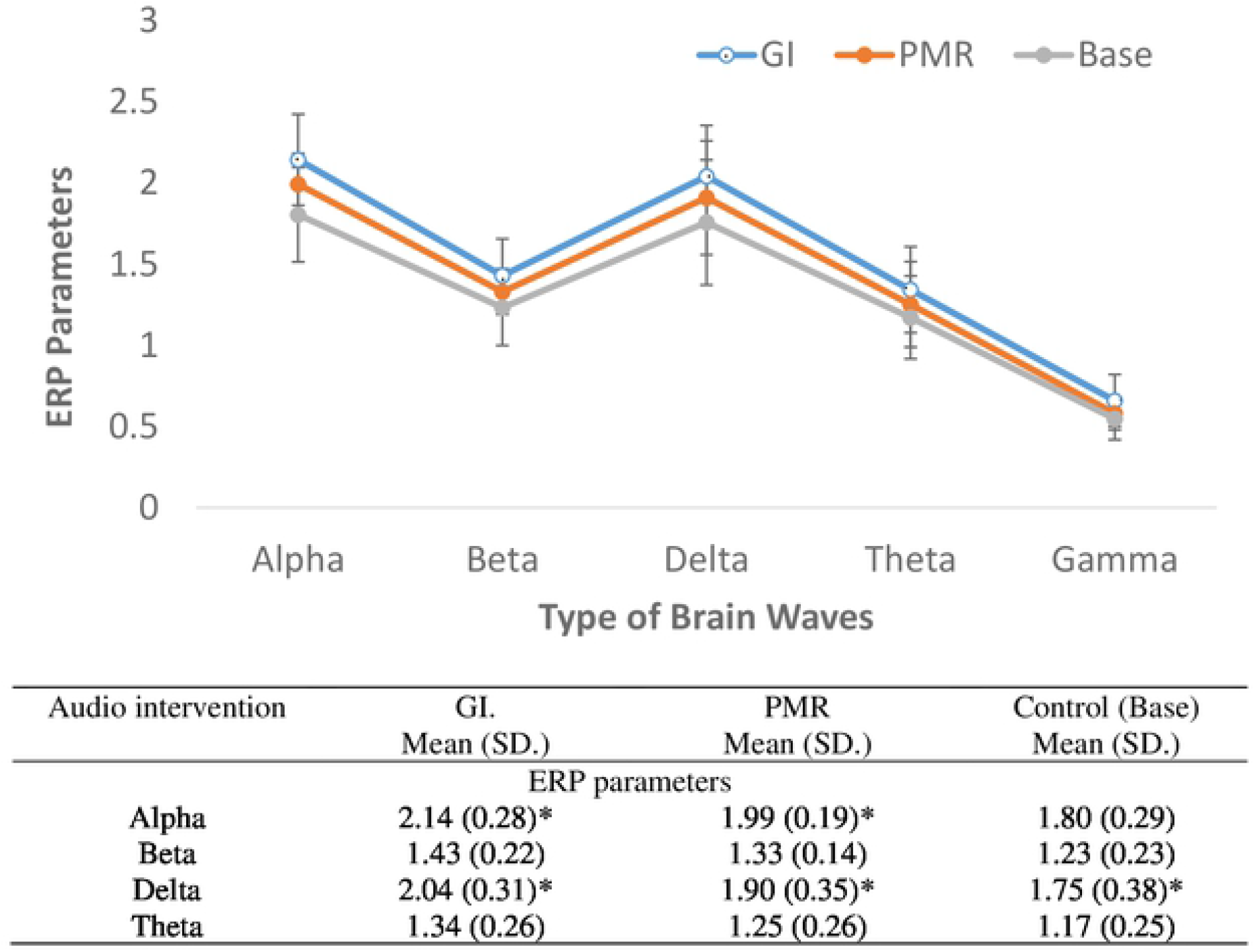
Reported values, pairwise comparison, and RM-ANOVA analysis of ERP parameters between guided imagery, progressive muscle relaxation and controls. Legend – GI: guided imagery, PMR: progressive muscle relaxation, ERP: brain event-related potentials. *Significant mean difference based on pairwise comparison in alpha waves between GI and PMR (p=0.029), delta waves between GI and PMR (p=0.029) and between PMR and control (p=0.014).

